# Genetically predicted serum vitamin D and COVID-19: a Mendelian randomization study

**DOI:** 10.1101/2021.01.29.21250759

**Authors:** Bonnie K Patchen, Andrew G Clark, Dana B Hancock, Nathan Gaddis, Patricia A Cassano

## Abstract

**Objectives:** To investigate causality of the association of serum vitamin D with the risk and severity of COVID-19 infection.

**Design:** Two-sample Mendelian randomization study.

**Setting:** Summary data from genome-wide analyses in the population-based UK Biobank and SUNLIGHT Consortium, applied to meta-analyzed results of genome-wide analyses in the COVID-19 Host Genetics Initiative.

**Participants:** 17,965 COVID-19 cases including 11,085 laboratory or physician confirmed cases, 7,885 hospitalized cases, and 4,336 severe respiratory cases, and 1,370,547 controls, primarily of European ancestry.

**Exposures:** Genetically predicted variation in serum vitamin D status, based on genome-wide significant single nucleotide polymorphisms (SNPs) associated with serum vitamin D or risk of vitamin D deficiency/insufficiency.

**Main outcome measures:** Susceptibility to and severity of COVID-19 infection, including severe respiratory infection and hospitalization.

**Results:** Mendelian randomization analysis, powered to detect moderate effects comparable to those seen in observational studies, provided little to no evidence for an effect of genetically predicted serum vitamin D on susceptibility to or severity of COVID-19 infection. Using SNPs in loci related to vitamin D metabolism as proxies for serum vitamin D concentration, the odds ratio for a standard deviation increase in serum vitamin D was 1.04 (95% confidence interval 0.92 to 1.18) for any COVID-19 infection versus population controls, 1.05 (0.84-1.31) for hospitalized COVID-19 versus population controls, 0.96 (0.64 to 1.43) for severe respiratory COVID-19 versus population controls, 1.15 (0.99 to 1.35) for COVID-19 positive versus COVID-19 negative, and 1.44 (0.75 to 2.78) for hospitalized COVID-19 versus non-hospitalized COVID-19. Results were similar in analyses that used all SNPs with genome-wide significant associations with serum vitamin D (i.e., including SNPs in loci with no known relationship to vitamin D metabolism) and in analyses using SNPs with genome-wide significant associations with risk of vitamin D deficiency or insufficiency.

**Conclusions:** These findings suggest that genetically predicted differences in long-term vitamin D nutritional status do not causally affect susceptibility to and severity of COVID-19 infection, and that associations observed in previous studies may have been driven by confounding. These results do not exclude the possibility of low-magnitude causal effects, nor do they preclude potential causal effects of acute responses to therapeutic doses of vitamin D. Future directions include extension of this work to non-European ancestry populations, and high-risk populations, for example persons with comorbid disease.

## INTRODUCTION

The COVID-19 pandemic has reached every corner of the globe and continues to spread. With more than 100 million cases and 2.0 million deaths globally at time of writing,^1^ identification of risk factors for susceptibility to SARS-CoV-2 infection and severity of COVID-19 is critical. Vitamin D nutritional status is a promising modifiable risk factor, and higher vitamin D is posited to reduce the risk of SARS-CoV-2 infection and the severity of the clinical course of COVID-19. Hypothesized vitamin D effects are biologically plausible given prior evidence that vitamin D upregulates innate and adaptive immunity to fight infection and reduce inflammation,^2^ is associated with a reduced risk of respiratory disease mortality,^3^ and enhances expression of ACE2, which is hypothesized to modulate the immune system response to SARS-CoV-2 infection.^4,5^ A recent *in vitro* study showed that vitamin D reduces viral load of nasal epithelial cells infected with SARS-CoV-2,^6^ and preliminary evidence from two small human trials suggested that vitamin D supplementation may help improve the prognosis of COVID-19 in infected individuals.^7,8^

Ecological and observational studies lend further support to the hypothesis that lower serum vitamin D concentrations are associated with both an increased risk of COVID-19 and an increased severity of the infection. Early in the pandemic, the prevalence of COVID-19 in European countries was negatively correlated with national averages for serum vitamin D concentration.^9^ COVID-19 mortality rates were positively correlated with latitude, a proxy for UV B exposure (higher latitude associates with lower UV B), which is required for synthesis of vitamin D in the body.^10,11^ More recent studies report associations of lower pre-pandemic serum vitamin D concentration or higher risk of vitamin D insufficiency with susceptibility to COVID-19 infection^12–14^ and with severity of COVID-19 infection in hospitalized patients as indicated by elevated biological markers of the cytokine storm, increased risk of intubation or use of supplemental oxygen, ICU admission, and death.^15–18^ The mounting evidence from observational studies and the known effects of vitamin D on the immune system contribute to speculation about whether a simple and immediate intervention like vitamin D supplementation might be effective in reducing risk of COVID-19 infection or severity, but sources of guidance for clinicians and the public cite a lack of evidence for a causal association.^19,20^ Further evidence is urgently needed.

The uncertainty about the causal association of vitamin D and COVID-19 arises because of the well-known association of serum vitamin D concentration with many known risk factors for COVID-19, including age, sex, body mass index (BMI), and race/ethnicity. The largest observational study of the UK Biobank data (N = 348,598 participants) reported little to no association of serum vitamin D with risk of COVID-19 infection in multivariate models adjusted for covariates.^21^ In the absence of large, rigorous randomized trials, it is challenging to determine whether other reported inverse associations of vitamin D and risk of COVID-19 are causal. Given the high global prevalence of vitamin D insufficiency, estimating the true effect of vitamin D on risk of COVID-19 infection and severity is important.

Mendelian randomization (MR), a study design to address causality, uses a form of instrumental variable analysis to improve causal inference and to address the biases inherent in observational studies. MR uses genetic variants that are associated with the exposure (i.e., serum vitamin D) as instrumental variables, or genetic instruments, that represent the long-term usual exposure. Following the laws of independent assortment, genetic variants are inherited from parent to offspring in a random and independent manner. An individual’s genotype therefore mimics the lifelong randomization of individuals into groups with different long-term serum vitamin D levels. Valid MR analysis depends on three key assumptions: (1) adequate strength and validity of the genetic instruments (i.e., genetic variants that underlie vitamin D metabolism), (2) independence of the genetic instruments from any confounders, and (3) absence of direct effects of the genetic instruments on the outcome of interest (i.e., COVID-19 status). MR addresses limitations in observational data including confounding, reverse causality, and measurement error, and supports triangulation on the evidence for causality when randomized trials are either impossible to conduct or currently unavailable. Genome-wide association (GWA) studies of serum vitamin D levels identified genetic variants that are robustly associated with serum vitamin D status in different populations and ancestries.^22–28^ Capitalizing on this finding, researchers successfully applied MR to study associations of vitamin D with many clinical outcomes, including diseases of immune system dysregulation and inflammation.^29–45^ We used MR to investigate causality of the relationship of serum vitamin D status with the risk and severity of COVID-19 infection.

## METHODS

### Two-sample Mendelian Randomization with Multiple Instruments

We estimated the effect of serum vitamin D levels on risk of COVID-19 infection and severity with two-sample MR.^46^ The associations of the genetic instruments with the exposure(s) and outcome(s) were estimated in separate, independent samples, then used to calculate the MR estimate of the effect of serum vitamin D status on COVID-19.^46^ We used summary data from a GWA study of serum vitamin D in the UK Biobank (sample 1)^22^ and genome-wide data for COVID-19 patients versus comparison groups from the COVID-19 Host Genetics Initiative (sample 2) (Figure 1).^47^ Because of overlap between the UK Biobank and the COVID-19 Host Genetics Initiative samples, which may bias MR results toward effects estimated from traditional observational studies,^48^ we replicated the analyses using summary GWAS data for serum vitamin D from the SUNLIGHT Consortium.^23^ We also performed sensitivity analyses to evaluate prior hypotheses about the direct effect of vitamin D deficiency or insufficiency on COVID-19 outcomes using summary data from a meta-analysis of associations of vitamin D SNPs with the dichotomous outcome of vitamin D deficiency versus sufficiency.^24^

**Figure 1).**
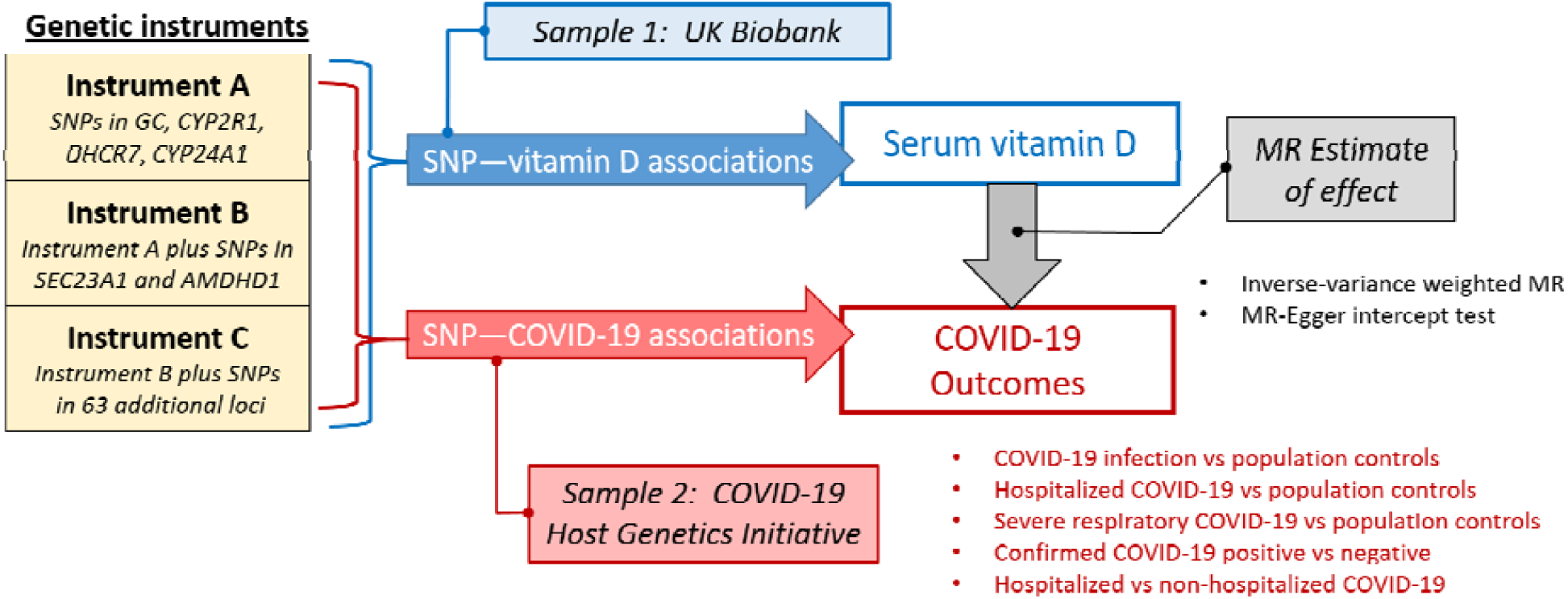
Schematic of two-sample MR study design.

### Vitamin D Genetic Instrument(s)

We constructed all genetic instruments for serum vitamin D levels based on independent genetic loci associated with serum vitamin D levels at genome-wide significance (p < 5×10^−8^). For the primary analysis, we used a biologically plausible genetic instrument (instrument A) consisting of SNPs in genetic loci that encode proteins in vitamin D transport and metabolism (Supplemental Figure 1), including the vitamin D binding protein (*GC*), 25 OH hydroxylase (*CYP2R1*), 7-dehydrocholesterol reductase (*DHCR7*), and 24-hydroxylase (*CYP24A1*). All SNPs included in Instrument A were associated with serum vitamin D at genome-wide significance in the UK Biobank and replicated in the SUNLIGHT Consortium.^22,23^

In secondary analyses, we expanded the genetic instrument to include additional genome-wide significant SNPs in loci with no known relation to vitamin D metabolism (Figure 1). The first expanded instrument (instrument B) included SNPs in two additional loci, Sec23 homolog A (*SEC23A*) and amidohydrolase domain containing 1 (*AMDHD1*), that were associated with serum vitamin D at genome-wide significance in both the UK Biobank and the SUNLIGHT Consortium. The second expanded instrument (instrument C) included SNPs in 63 additional loci associated with serum vitamin D at genome-wide significance in the UK Biobank (Supplemental Table 1).

Finally, we constructed separate genetic instruments for risk of vitamin D deficiency and insufficiency (defined as serum vitamin D < 50 nmol/L and <75 nmol/L, respectively) based on meta-analysis results of vitamin D SNP—risk of vitamin D deficiency/insufficiency associations in the SUNLIGHT Consortium.^24^ The genome-wide significant SNPs contributing to the vitamin D deficiency and insufficiency instruments were all in loci related to vitamin D metabolism, including *GC, DHCR7*, and *CYP2R1*.

### SNP—Vitamin D Associations

We extracted summary data, including beta-coefficients, standard errors, and effect alleles and their frequencies for each SNP contributing to the genetic instruments. We primarily used summary data from a UK Biobank GWAS of serum vitamin D in participants of “white British” ancestry as defined by genotype principal component analysis (N = 401,460)^22^ and replicated the analyses using summary data from a SUNLIGHT Consortium meta-analysis GWAS of serum vitamin D (31 European ancestry cohorts, N = 79,366).^23^ For sensitivity analyses, we used data from a SUNLIGHT Consortium meta-analysis of associations of vitamin D SNPs with risk of vitamin D deficiency/insufficiency (four European ancestry cohorts, N = 16,905).^24^

Details of the UK Biobank and SUNLIGHT Consortia GWA studies of serum vitamin D and the candidate SNP—vitamin D deficiency/insufficiency meta-analysis are described elsewhere.^22–24^ Briefly, in the UK Biobank GWAS, serum vitamin D data were log-transformed and standardized to a mean of 0 and standard deviation of 1, and SNP–vitamin D association models were adjusted for age, sex, season of vitamin D measurement, vitamin D supplementation, genotype batch, genotype array, and assessment center (as a proxy for latitude). In the SUNLIGHT Consortium meta-analysis GWAS of serum vitamin D, serum vitamin D data were log-transformed and SNP–vitamin D association models were adjusted for age, sex, BMI, month of sample collection, cohort-specific variables such as geographical location and assay batch, and genotype principal components.^22^ In the SUNLIGHT Consortium meta-analysis of SNP—risk of vitamin D deficiency/insufficiency associations, models were adjusted for age, sex, BMI, and season, and genomic control was applied to control for population stratification.^24^

### Vitamin D Instrument Strength

We assessed the strength and validity of the genetic instruments for vitamin D by calculating the F-statistic,^49^ according to the method described by Burgess and colleagues for two-sample MR.^48^ First, we calculated the variance in vitamin D explained by each SNP (R^2^ _snp_) using the equation R^2^ _snp_ =2 α ^2^ MAF(1-MAF), where α = SNP–serum vitamin D association and MAF = minor allele frequency. Next, we calculated the F-statistic for each of the instruments, using the equation F=(N-K-1)/K x (R^2^_instrument_)/(1-R^2^ _instrument_), where N = sample size, K = number of SNPs contributing to the genetic instrument, and R^2^ _instrument_= sum of R^2^ _snp_ across SNPs contributing to the instrument. We based all calculations on GWAS data from the UK Biobank^22^ and calculated the F-statistic for sample sizes of 10,000 and 1.3 million individuals, the approximate minimum and maximum sample sizes for the COVID-19 outcomes. We considered the MR standard of F-statistic >10 as an indicator of instrument strength.^49^

### SNP—COVID Associations

We used genome-wide data for COVID-19 cases and comparison groups from the COVID-19 Host Genetics Initiative, which is described elsewhere.^47^ Briefly, the initiative comprises a global effort to study genetic contributions to variability in COVID-19 infection and severity. Contributing studies performed genome-wide analyses for COVID-19 case and comparison groups following an analysis plan developed by the initiative. Studies with multiple ancestries performed stratified analyses, and all analyses were adjusted for sex, age, age,^2^ sex × age interaction, and genotype principal components. Results were summarized across studies and ancestries by meta-analysis.

The publicly available summary results were downloaded from the COVID-19 Host Genetics Initiative’s fourth data freeze and meta-analysis (https://www.covid19hg.org/results/), which were released on October 20, 2020. We considered the following five COVID-19 case versus control comparisons, using definitions provided by the initiative: (1) any COVID-19 versus population controls, (2) hospitalized COVID-19 versus population controls, (3) very severe respiratory confirmed COVID-19 versus population controls, (4) any COVID-19 versus confirmed COVID-19 negative, and (5) hospitalized versus non-hospitalized COVID-19. For each COVID-19 “case” versus comparison group, we extracted summary data, including beta-coefficients, standard errors, effect alleles, and effect allele frequencies corresponding to the SNPs included in the vitamin D genetic instruments. For SNPs that were not available in the public data, we identified proxy SNPs based on linkage disequilibrium (LD), using the standard MR threshold for LD proxies of r^2^ > 0.8.^50^ All LD proxy SNPs were defined using 1000 Genomes European (CEU and GBR) sample data and identified through LDlink.^51^ Case and comparison group definitions, sample sizes, and ancestry distributions for the different comparisons are summarized in Table 1. Studies contributing to these analyses are listed in Supplemental Table 1.

**Table 1.** Sample Description for COVID-19 case and comparison groups.

| Comparison | Case definition | Comparison group definition | Sample size | Ancestry |
| --- | --- | --- | --- | --- |
| Any COVID-19 case vs. population controls | laboratory/physician confirmed OR self-reported positive | Everyone that is not a case (e.g., the population) | 17,965 cases, 1,370,547 controls | 93.6% EUR<br>3.1% EAS/SAS<br>1.8% AFR<br>1% ARAB<br><1% AMR<br><1% HIS |
| Hospitalized COVID-19 cases vs. population controls | laboratory/physician confirmed AND hospitalized for COVID-19 | Everyone that is not a case (e.g., the population) | 7,885 cases, 961,804 controls | 93.7% EUR<br>3.5% EAS/SAS<br>1.1% AFR<br>1.4% ARAB<br><1% AMR<br><1% HIS |
| Severe respiratory COVID-19 cases vs. population controls | laboratory confirmed AND hospitalized AND respiratory support or death | Everyone that is not a case (e.g., the population) | 4,336 cases, 623,902 controls | 99.6% EUR<br><1% AMR |
| COVID-19 positive cases vs. COVID-19 negative controls | laboratory/physician confirmed OR self-reported positive | laboratory confirmed OR self-reported negative | 11,085 cases, 116,794 controls | 86.5% EUR<br><1% EAS/SAS<br>9.6% AFR<br>3.2% HIS |
| Hospitalized vs. nonhospitalized COVID-19 | laboratory/physician confirmed AND hospitalized for COVID-19 | Laboratory or physician confirmed COVID-19 | 2,430 hospitalized<br>8,478 nonhospitalized | 75.3% EUR<br><1% EAS/SAS<br>12.6% AFR<br>6.4% ARAB<br>4.7% HIS |

AFR = African; ARAB = Arabic; AMR = Ad Mixed American; EAS/SAS = East Asian/South Asian; EUR = European; HIS = Hispanic

Summary data were available for mixed ancestry analyses for all outcomes. For the outcomes of COVID-19 versus population controls and hospitalized COVID-19 versus population controls, summary data from analyses limited to European ancestry participants were also available. For these outcomes, we extracted summary data from both the mixed ancestry and the European ancestry analyses for comparison and to address differences in ancestry composition between vitamin D GWAS and COVID-19 Host Genetics Initiative sample populations.

### Stratification of SNP—Vitamin D Associations by COVID-19 Risk Factors

Our study design assumes that the potential effect of vitamin D nutritional status on susceptibility to and severity of COVID-19 infection is the same across different COVID-19 risk categories, including ancestry, age, sex, BMI, and smoking status. MR analysis under this assumption requires the genetic instruments for vitamin D to be robust across the COVID-19 risk subgroups. To address this assumption, we conducted stratified analyses of the SNP-vitamin D associations for all SNPs contributing to the genetic instruments using data from the UK Biobank. UK Biobank Resource data—including serum vitamin D levels, vitamin D supplementation, covariates and stratification variables, and imputed genotype dosages with reference to Haplotype Reference Consortium and UK10K haplotype resource—were obtained under Application Number 24603. The Institutional Review Board at RTI International approved the UK Biobank data use and analysis.

The SNP–vitamin D associations were estimated stratified by sex (male vs. female), age group (40-50 vs. 50-60 vs. 60-70), BMI category (18.5-25 vs. 25-30 vs. 30-40 vs. >40), and smoking status (never vs. former vs. current smokers). Analyses were performed using the GENESIS R/ Bioconductor package,^52^ separately for European ancestry (N = 421,407) and African ancestry (N = 7,859) participants, as classified via analysis with STRUCTURE^53^ in comparison to the CEU, YRI, and CHB 1000 Genomes populations.^54,55^ Statistical models resembled those described for SNP–vitamin D associations in UK Biobank,^22^ except that genotype batch was not included as a covariate and vitamin D levels were not log-transformed.

### Statistical Analysis for Two-Sample MR

We performed two-sample MR using the inverse variance weighted (IVW) method, which assumes that pleiotropy is either non-existent or balanced (i.e., any associations of genetic instrument SNPs with phenotypes other than vitamin D are randomly positive and negative such that the mean pleiotropic effect is zero).^56^ We used the MR-Egger intercept test to check for evidence of directional pleiotropy, and performed sensitivity analyses using the MR-Egger, weighted-median and weighted-mode methods.^57–59^ The MR-Egger method relaxes the assumption of no directional pleiotropy and corrects for pleiotropy-induced bias by allowing for a nonzero intercept.^56,57^ The weighted-median method assumes that at least 50% of the weight of the genetic instrument comes from nonpleiotropic SNPs and produces an unbiased estimate so long as less than 50% of SNPs have pleiotropic effects.^58^ The weighted-mode method assumes that the most common (modal) SNP–vitamin D effect is the true effect and therefore allows for the inclusion of pleotropic SNPs without biasing the MR estimate.^59^ Where evidence of directional pleiotropy existed (MR-Egger intercept test P-value <0.05) we prioritized the MR estimates from the MR-Egger, mode-weighted, and median-weighted analyses, otherwise we prioritized the estimates from the IVW method. Results for our primary analysis are presented as odds ratios and 95% confidence intervals (OR, 95% CI) for COVID-19 case versus comparator per standard deviation increase in log-transformed serum vitamin D. The threshold for statistical significance was set at a P-value < 0.05. All analyses were performed using R Studio version 1.2.1335 and the “TwoSampleMR” R package (https://github.com/MRCIEU/TwoSampleMR)^50^.

We estimated potential bias under the null due to sample overlap between the UK Biobank vitamin D GWAS and the COVID-19 Host Genetics Initiative using estimates of the vitamin D–COVID-19 associations from observational studies, the percentage of sample overlap, and the relative bias (reciprocal of the F-statistic), as previously described.^48^ We compared SNP-vitamin D associations stratified by sex, age group, BMI category, and smoking status with Pearson’s correlation coefficients.

We calculated the minimum odds ratio detectable at 80% power for each COVID-19 outcome using the “mRnd” tool (https://shiny.cnsgenomics.com/mRnd/).^60^ Based on heritability estimates from the UK Biobank and SUNLIGHT Consortium vitamin D GWA studies,^23,61^ we assumed the genetic instruments accounted for 3% of the variance in serum vitamin D for all power calculations.

### Patient and Public Involvement

We used publicly available data from several consortia including the UK Biobank, the SUNLIGHT Consortium and the COVID-19 Host Genetics Initiative in this study. These consortia were not involved in any stage of the design and conduct of this research, nor were they asked to advise on interpretation or writing up of results. No patients were involved in any stage of the research process. There are no plans to disseminate the results of the research to study participants or the relevant patient community.

## RESULTS

### Genetic Instruments for Vitamin D

Characteristics of the genetic instruments for serum vitamin D, including contributing SNPs, SNP–vitamin D associations in the UK Biobank, and F-statistics, indicate that instruments

A and B clearly exceed the MR standard for instrument strength of F-statistic > 10 across the range of COVID-19 outcome sample sizes (Table 2, Supplemental Table 2). The F-statistics for instruments A, B, and C were respectively 53, 37, and 5 for the minimum sample size of 10,000 participants and 6,915, 4,746, and 600 for the maximum sample size of 1.3 million participants. Instrument A, which was limited to SNPs in loci related to vitamin D metabolism (Supplementary Figure 1), had the largest F-statistics across all sample sizes, reflecting the strength of the SNP–vitamin D associations for the SNPs in loci related to the vitamin D metabolic pathway.

**Table 2.** Characteristics of vitamin D instruments and association with measured vitamin D status in UK Biobank (N=401,460, Source: Manousaki et al., 2020)

| Loci | SNP | EA | NEA | EAF | Beta-vitD | P-vitD | F-statistic |  |
| --- | --- | --- | --- | --- | --- | --- | --- | --- |
|  |  |  |  |  |  |  | N=10,000 | N=1,300,000 |
| Instrument A |  |  |  |  |  |  | 53 | 6,915 |
| GC | rs11723621 | G | A | 0.29 | -0.19 | 2.9E-1689 | 147 | 19,072 |
| CYP2R1 | rs10832289 | T | A | 0.41 | -0.07 | 2.03E-266 | 23 | 4,597 |
| DHCR7 | rs12803256 | G | A | 0.78 | 0.10 | 1.3E-378 | 35 | 715 |
| CYP24A1 | rs6127099 | T | A | 0.28 | -0.04 | 9.30E-62 | 5 | 6,915 |
| Instrument B |  |  |  |  |  |  | 37 | 4,746 |
| SEC23A | rs8018720 | C | G | 0.82 | -0.03 | 4.04E-36 | 3 | 391 |
| AMDHD1 | rs10859995 | C | T | 0.58 | -0.04 | 7.03E-89 | 8 | 983 |
| Instrument C* |  |  |  |  |  |  | 5 | 600 |

\*Characteristics of all SNPs in instrument C are shown in Supplemental Table 1
Beta-vitD = beta coefficient for association of an additional effect allele with a SD change in serum vitamin D in the log scale; EA = effect allele; EAF = effect allele frequency; F-statistic = measure of instrument strength, calculated as described in the methods, shown for approximate minimum and maximum sample sizes across the COVID-19 outcomes; NEA = non-effect allele; P-vitD = P-value for the beta coefficient; SNP = single nucleotide polymorphism

### COVID-19 Case-Comparison Groups and Power Calculations

We considered multiple case definitions, sample sizes, and ancestral compositions for the COVID-19 case-control comparisons in our analysis (Table 1). Sample sizes across the analyses ranged from just over 10,000 (for hospitalized COVID-19 vs. non-hospitalized COVID-19) to over 1.3 million (for COVID-19 vs. population). The majority (>75%) of participants were of European ancestry. Non-European ancestry participants made up a much smaller fraction of the total sample.

Based on the sample sizes across the COVID-19 case-control comparisons, and assuming our genetic instruments explained 3% of the variance in serum vitamin D, we calculated that this study had 80% power to detect moderate effect sizes comparable to those seen in observational studies. The minimum detectable ORs for a standard deviation increase in serum vitamin D ranged from 0.66 (hospitalized COVID-19 vs. non-hospitalized COVID-19) to 0.86 (COVID-19 vs. population; Supplemental Table 3).

### MR Estimates for Serum Vitamin D Effect on COVID-19 Outcomes

MR estimates (odds ratios and 95% confidence intervals) for the effect of genetically predicted serum vitamin D on risk of COVID-19 outcomes, calculated separately for instruments A, B, and C, are shown in Figure 2 and Table 3. MR scatter plots and forest plots further illustrating these results are shown in Supplementary Figure 2. MR-Egger intercept tests for pleiotropy were not statistically significant (MR-Egger intercept p-values > 0.1) for all instruments and all outcomes (Table 3). Thus, we prioritized the MR estimates from the IVW analyses and found little to no evidence for an effect of vitamin D on risk of any COVID-19 outcome considered. For instrument A, using SNPs in loci related to vitamin D metabolism, the odds ratio for a standard deviation increase in serum vitamin D was 1.04 (95% confidence interval 0.92 to 1.18) for any COVID-19 infection versus population controls, 1.05 (0.84-1.31) for hospitalized COVID-19 versus population controls, 0.96 (0.64 to 1.43) for severe respiratory COVID-19 versus population controls, 1.15 (0.99 to 1.35) for COVID-19 positive versus COVID-19 negative, and 1.44 (0.75 to 2.78) for hospitalized COVID-19 versus non-hospitalized COVID-19 (p-values >0.1 for all comparisons). Results were similar for instruments B and C. Results of MR analyses limited to European ancestry participants, which we were able to perform for the outcomes of COVID-19 versus population and hospitalized COVID-19 versus population, were also non-significant (Supplementary Figure 3).

**Table 3.**
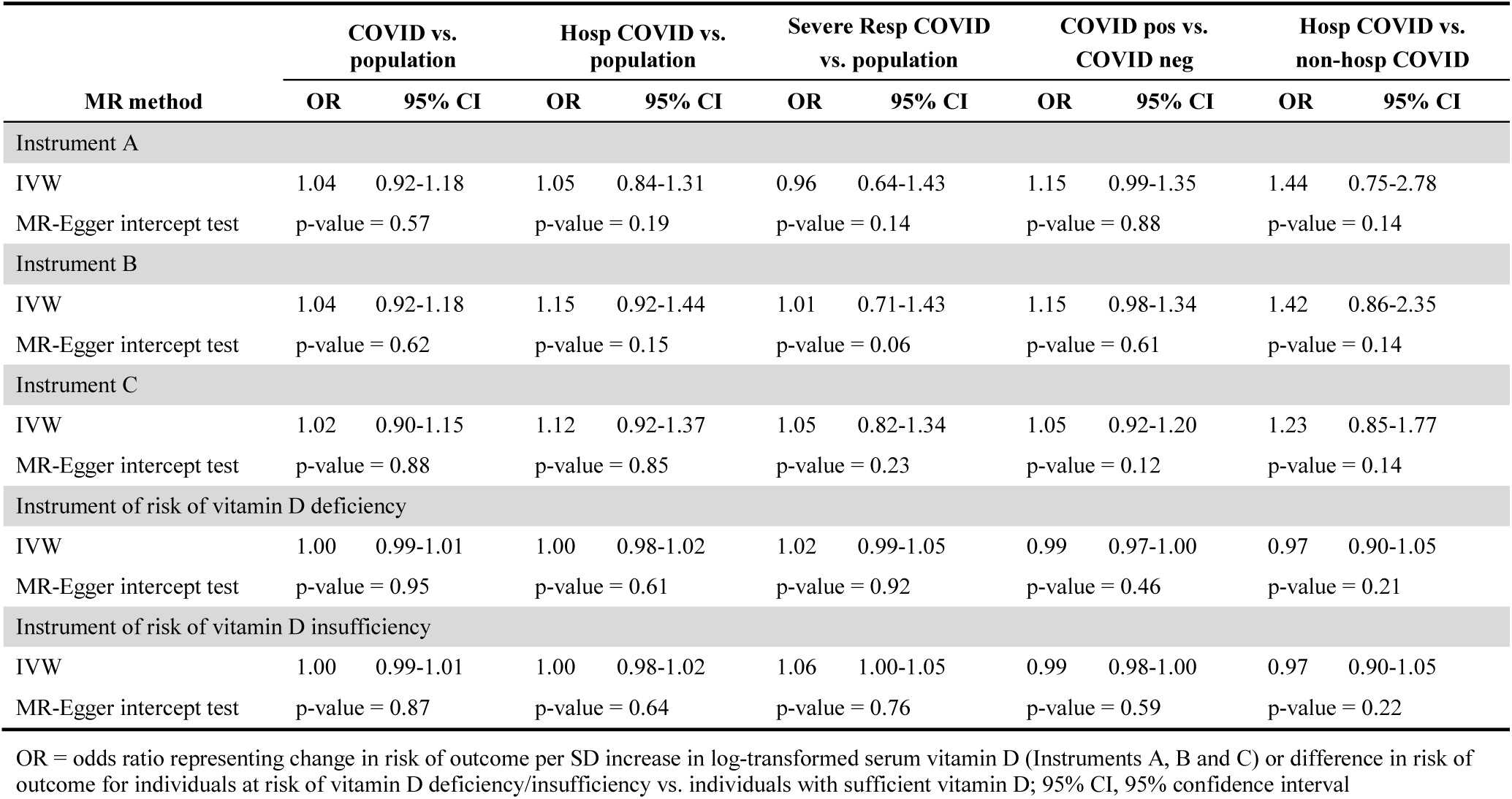
Mendelian Randomization (MR) estimates of effect of vitamin D on COVID-19 outcomes.

**Figure 2).**
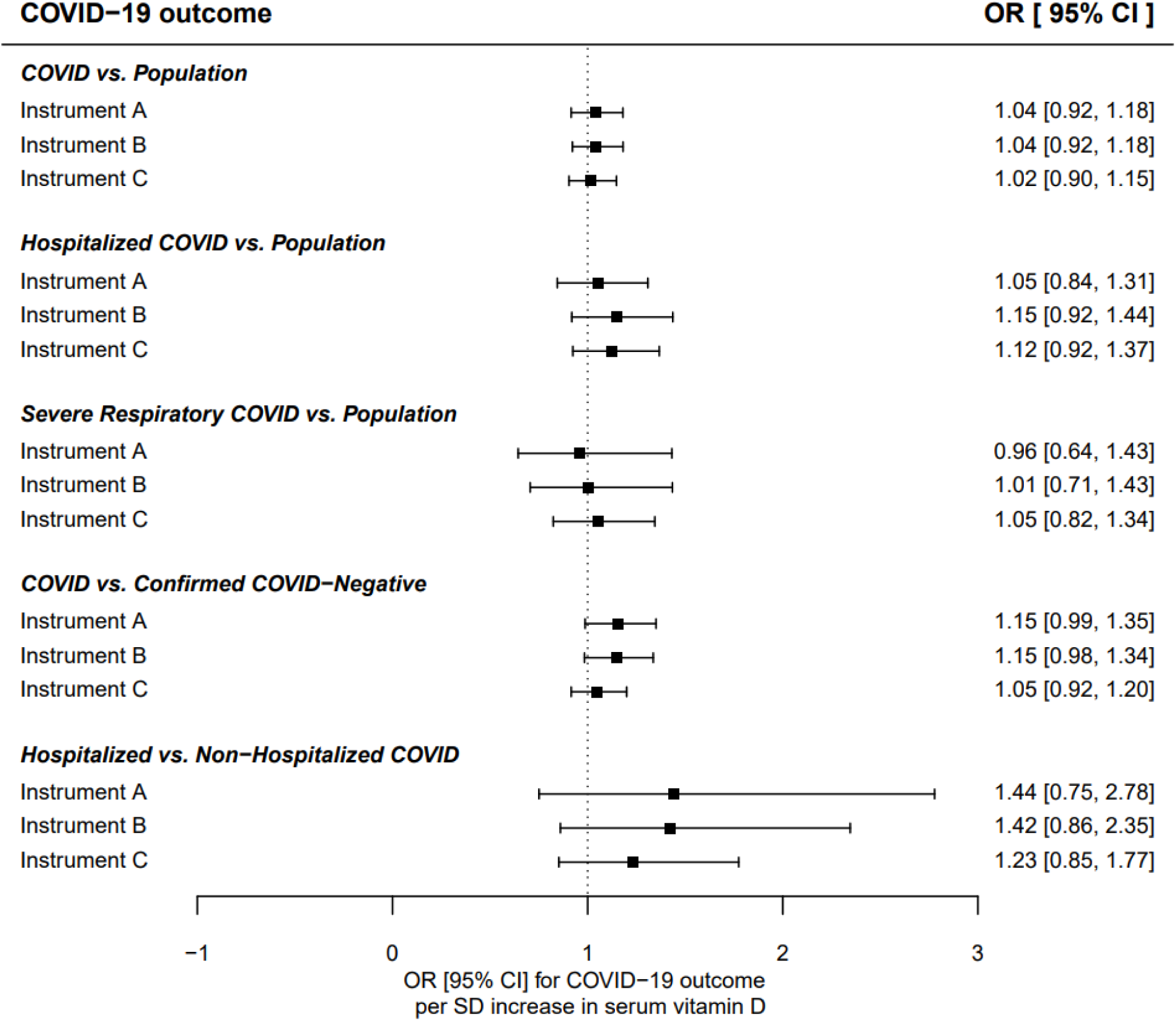
Inverse variance weighted MR estimates for effect of serum vitamin D on risk of COVID-19 outcomes.

**Figure 3).**
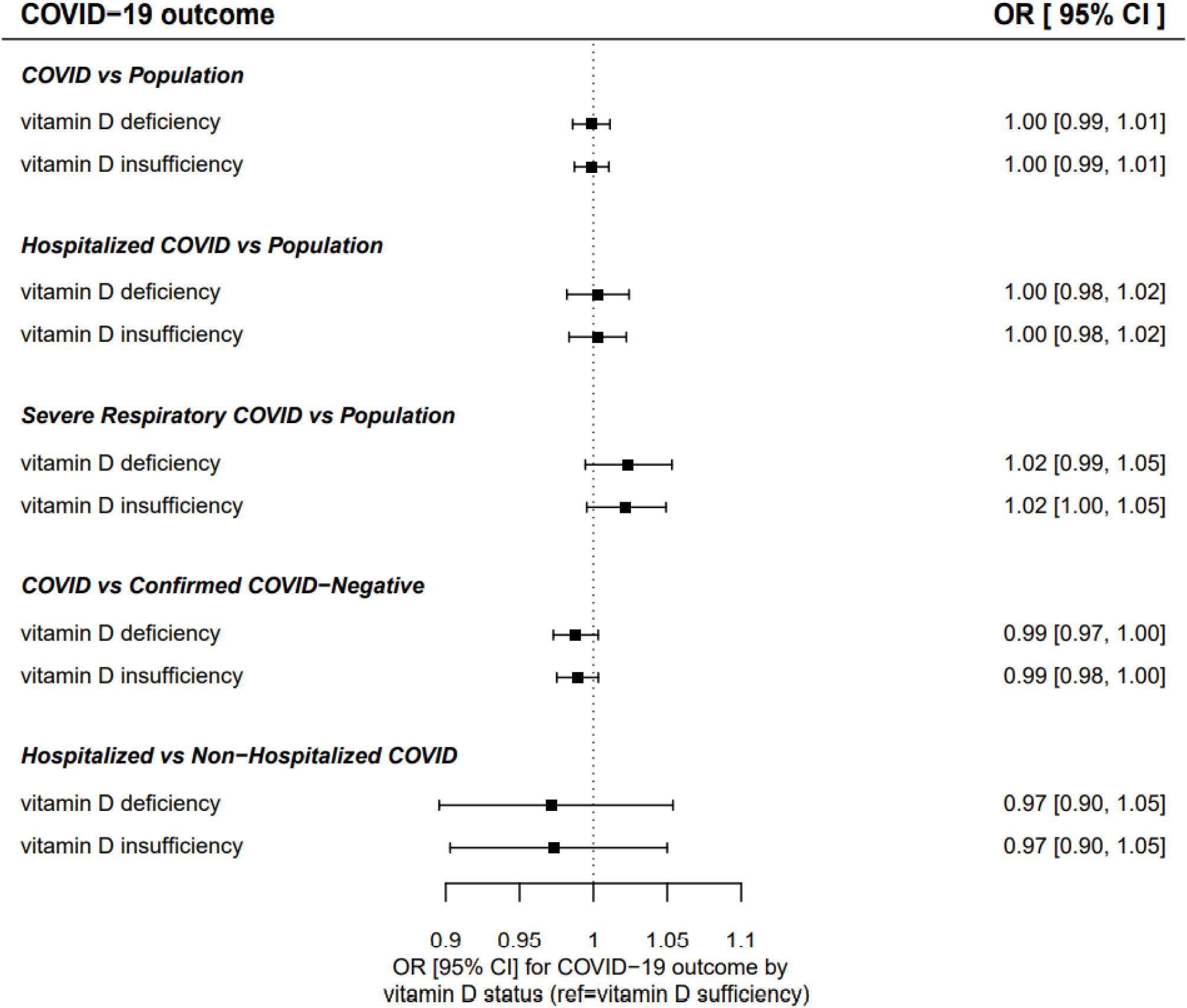
Inverse variance weighted MR estimates for effect of vitamin D insufficiency or deficiency on risk of COVID-19 outcomes.

For all outcomes, the MR-Egger intercept test showed little to no evidence of pleiotropy (Table 3). Nevertheless, we performed sensitivity analysis to explore possible pleiotropy using the MR-Egger, weighted-median, and weighted-mode MR methods, and this analysis showed a *direct* association of serum vitamin D with risk of hospitalized COVID-19 versus non-hospitalized COVID-19 (Supplemental Figure 4). For this outcome, MR estimates from the MR-Egger and mode-weighted analyses found that chronically *higher* vitamin D levels increase the odds of hospitalization in COVID-19-infected participants (OR [95% CI] for instrument A: 1.83 [1.12, 2.97] for mode-weighted MR and 3.69 [1.53, 8.93] for MR-Egger). MR estimates from the MR-Egger, mode- and median-weighted MR analyses were non-significant for all other outcomes, consistent with results from the IVW analysis.

Estimations of bias because of participant overlap between the UK Biobank and the COVID-19 Host Genetics Initiative were negligible. For the outcome with maximum participant overlap (58% for severe respiratory COVID-19 vs. population), we estimated the bias for instruments A, B, and C to be 0.1%, 0.2%, and 1.0% respectively. We confirmed the negligible effect of participant overlap in our study by repeating the analyses using summary data for GWAS significant SNP–vitamin D associations from the SUNLIGHT consortium in place of UK Biobank data (Supplementary Table 4). Results using the SUNLIGHT Consortium data were consistent with results using the UK Biobank data; there was no evidence of pleiotropy for any outcome (MR-Egger intercept p-value >0.1), and IVW MR estimates for the effect of genetically predicted serum vitamin D on risk of COVID-19 outcomes were similar in magnitude and direction and were not statistically significant (Supplementary Table 4).

### MR Estimates for Vitamin D Deficiency/Insufficiency and COVID-19 Outcomes

A further sensitivity analysis evaluated the effect of genetically predicted vitamin D deficiency and insufficiency on COVID-19 outcomes (Table 3, Figure 4). MR-Egger intercept tests for pleiotropy were all non-significant (p-value > 0.1), and the IVW MR estimates did not support an effect of long-term vitamin D deficiency or insufficiency on the risk of any of the COVID-19 outcomes evaluated (Table 3, Figure 4).

### Covariate Effects on SNP-Vitamin D Associations

Stratified analyses of SNP-vitamin D associations evaluated the validity of the genetic instruments across known COVID-19 risk factors, including ancestry, age, sex, BMI, and smoking status. Correlations of age-, sex-, BMI-, and smoking status-specific SNP–vitamin D associations are shown separately for European (N = 421,407) and African (N = 7,859) ancestry participants in Supplemental Figure 5. For European ancestry participants, subgroup-specific SNP–vitamin D associations were strongly correlated across age, sex, BMI category, and smoking status (Pearson’s correlation coefficients ranging from 0.90 to 0.99), supporting the use of the instrument in the general European ancestry population. For African ancestry participants, the correlations among subgroup-specific SNP–vitamin D associations were less clear (Pearson’s correlation coefficients ranged from −0.95 to 0.72), suggesting that optimization of the genetic instruments for the African ancestry population may be necessary. Given the small sample sizes in the African ancestry subgroups, further research is needed to validate the instruments and their use in African and other non-European Ancestry populations.

## DISCUSSION

We used summary data from genome-wide analysis in the UK Biobank, SUNLIGHT Consortium, and COVID-19 Host Genetics Initiative to conduct the first MR study of serum vitamin D and COVID-19 outcomes. Although our study was powered to detect effects comparable to those seen in observational studies of vitamin D and COVID-19, we found little to no evidence of an effect of genetically predicted serum vitamin D levels on COVID-19 outcomes, including risk of COVID-19 infection, severe respiratory infection, and hospitalization. Our results were robust to multiple genetic instruments for vitamin D and were replicated using SNP–vitamin D association data from two independent samples. Results from sensitivity analyses evaluating the effect of genetically predicted vitamin D deficiency or insufficiency on risk of COVID-19 outcomes were similarly null. In summary, our results suggest that long-term usual vitamin D nutritional status does not have a causal effect on susceptibility to COVID-19 infection and its severity.

Our results are consistent with findings from the largest observational study to date of serum vitamin D and COVID-19 infection,^21^ which used UK Biobank data on approximately 350,000 White, Black, and South Asian participants, and found associations of pre-pandemic vitamin D levels with COVID-19 infection in univariate models, but not in models adjusting for confounding by known COVID-19 risk factors, including age, sex, race/ethnicity, BMI, socioeconomic status, smoking status, diabetes, blood pressure, chronic illness, and disability. We used the MR approach as an alternative way to address potential confounding. We instrumented long-term serum vitamin D status with genetic variants that are unlikely to have direct associations with known risk factors for COVID-19 infection. Specifically, the genetic instrument for our primary analysis (instrument A) was limited to SNPs in loci with functional links to serum vitamin D, including *GC, DHCR7, CYP2R1, and CYP24A1*. With this approach, we saw no evidence for an effect of genetically predicted (i.e., unconfounded) variation in serum vitamin D on risk and severity of COVID-19 infection.

These results are contrary to findings from several smaller observational studies showing an association of lower vitamin D levels with higher risk of testing positive for COVID-19 in the general population^12–14^ and higher risk of COVID-19 related hospitalization, ICU admission, intubation and death among infected individuals.^15–18^ However, these studies were inconsistent in the covariates included in statistical models, and in several studies estimates of vitamin D associations with susceptibility to and severity of COVID-19 infection were attenuated with covariate adjustment, suggesting the observed effects could be the result of residual confounding. Additionally, many of these studies were case-control or cross-sectional studies, making it difficult to rule out the possibility of reverse causality (i.e., that COVID-19 infection or its symptoms could lead to lowering of serum vitamin D). The MR approach addresses key limitations of these studies, including confounding and reverse causality, and is recognized as a method for improving causal inference in epidemiology.

This study has many strengths. We used summary data from the largest GWA studies of serum vitamin D to maximize the strength and validity of the genetic instruments for vitamin D and leveraged publicly available data on nearly 1.4 million participants, including 17,965 COVID-19 cases, from 38 study cohorts in the COVID-19 Host Genetics Initiative. We addressed potential pleiotropy of the genetic instruments, which if present would limit the interpretation of the MR results, by comparing results across the biologically plausible and expanded instruments, testing for evidence of pleiotropy with the MR-Egger intercept test, and performing sensitivity analysis with alternative MR methods that address pleiotropy. We explored potential threshold effects using genetic instruments for vitamin D deficiency and insufficiency. Our findings were robust across all exploration, and, in triangulation with results of the largest to date observational study of vitamin D and COVID-19, the findings help to clarify the nature of the association of vitamin D with COVID-19 outcomes.

As in any MR study, the valid interpretation of our results rests on the MR assumptions of adequate strength of the genetic instruments for vitamin D and absence of direct effects of the instruments on COVID-19 outcomes and potential confounders (i.e., pleiotropy). Our instruments easily met the MR standard for instrument strength (F-statistic >10) and we found no evidence of pleiotropy from the MR-Egger intercept test. Our results were consistent across the biologically plausible and expanded genetic instruments and largely similar in sensitivity analysis using MR methods designed to address pleiotropy. While the findings in our study are unlikely to be biased by pleiotropy, exhaustive tests for pleiotropy are challenging, making the MR assumptions difficult to fully verify.

We used a two-sample MR approach, using estimates of associations of the genetic instruments with vitamin D and with COVID-19 outcomes from two separate samples. This approach maximizes sample size and reduces the likelihood of bias toward estimates from observational studies^46^ but requires that the two samples are drawn from similar populations. In our study, the summary data for associations of the genetic instruments with vitamin D were from analyses in European ancestry participants, while the data for associations of the genetic instruments with COVID-19 outcomes were from analyses including up to 25% non-European ancestry participants. Repeating the analysis with a subgroup of European-only participants for two outcomes, COVID-19 versus population controls and hospitalized COVID-19 versus population controls, indicated minimal impact of ancestral differences on MR estimates.

However, ancestry-specific data were not available for COVID-19 outcomes with larger proportions of non-European ancestry participants, and the impact of ancestral differences could not be fully explored. In sensitivity analyses using alternative MR methods that assume some pleiotropy, findings indicated an association of *higher* vitamin D levels with *higher* risk of hospitalization among COVID-19-infected individuals, the outcome with the highest proportion of non-European ancestry participants. This finding could be driven by racial/ethnic disparities in access to health care (i.e., hospitalized COVID-19 individuals may be more likely of European ancestry and therefore have higher vitamin D status). We also found evidence for racial differences in risk-factor stratified SNP–vitamin D associations. Our results suggest that SNP– vitamin D associations are strongly correlated across sex, age, BMI, and smoking subgroups for individuals of European ancestry, but not for individuals of African ancestry, although because of small sample sizes for the African ancestry subgroups we are unable to draw firm conclusions. Further research using data from ancestry-specific GWA studies of vitamin D and COVID-19 outcomes is warranted to further investigate the impact of race/ethnicity on the vitamin D– COVID-19 relationship.

This study was not designed to evaluate the effect of acute changes in vitamin D status (i.e., from supplementation) on prevention or treatment of COVID-19. One randomized trial and one quasi-experimental trial have provided evidence for a positive effect of vitamin D supplementation around time of diagnosis on COVID-19 prognosis in infected individuals. The doses of vitamin D administered in these studies could result in an acute change in the availability of vitamin D, which may support the immune system’s response to the virus, mitigate acute lung injury, and contribute to improved prognosis. Our results, which pertain to long-term vitamin D status, do not preclude the possibility that therapeutic doses of vitamin D may be effective in preventing or treating COVID-19 infection. Larger randomized trials using diverse sample populations are needed to investigate the potential use of therapeutic doses of vitamin D supplementation for COVID-19 prevention and treatment.

In conclusion, we used two-sample MR to study the associations of vitamin D with COVID-19 outcomes to address limitations of existing observational studies, including confounding and reverse causality. We found no evidence for an effect of genetically predicted variation in serum vitamin D on risk or severity of COVID-19 infection. Our findings suggest that chronic differences in serum vitamin D do not have a causal effect on susceptibility to COVID-19 infection or severity of COVID-19 among those infected, and that associations observed in previous studies may have been driven by confounding. Future directions of this work include extension of the MR approach to non-European ancestry populations, and investigation of potential modification of genetically predicted vitamin D effects on COVID-19 risk and severity by COVID-19 risk factors, including race/ethnicity, age, sex, and BMI. Randomized trials are needed to inform the question of whether acute changes in vitamin D levels (i.e., from supplementation) are efficacious in COVID-19 prevention and treatment across diverse populations.

## SUMMARY BOX

### What is already known on this topic

- Observational studies report that lower vitamin D levels are associated with increased risk and severity of COVID-19
- Known risk factors for COVID-19 are associated with lower vitamin D levels in blood
- It is unknown whether observed associations of vitamin D and COVID-19 are causal

### What this study adds

- This study found no evidence for associations of genetically predicted long-term vitamin D levels with risk and severity of COVID-19 infection
- The findings do not support a causal effect of long-term usual vitamin D nutritional status on COVID-19 infection and its severity
- These findings have implications for the potential effectiveness of low level vitamin D supplementation as a strategy for COVID-19 prevention and treatment, but do not address the therapeutic use of vitamin D in acute disease

## Supporting information

Reporting Checklist

## Data Availability

All data used for this analysis are publicly available. Code implementing the MR analysis is available upon request from the corresponding author.

https://www.covid19hg.org/results/

## FOOTNOTES

### Contributor and Guarantor Confirmation

The study was designed and conceived by BKP, PAC and AGC. The MR analyses were performed by BKP in close consultation with PAC. The follow-up stratified analyses of SNP—vitamin D associations were performed by NG. The manuscript was drafted by BKP and PAC, and edited with input from AGC, DBH and NG. All authors reviewed and contributed to the discussion of findings and the crafting of the manuscript and gave final approval to the version submitted for publication. PAC is the guarantor.

### Ethics approval

The main analysis conducted in this study used publicly available summary data and did not require ethical approval. The use of UK Biobank data for the stratified analysisof SNP—vitamin D associations by COVID-19 risk factors was approved by the Institutional Review Board at RTI International.

### Transparency

PAC is the corresponding author and guarantor for this paper and affirms that the manuscript is an honest, accurate, and transparent account of the study being reported; that no important aspects of the study have been omitted; and that any discrepancies from the study as originally planned have been explained.

### Copyright

The Corresponding Author has the right to grant on behalf of all authors and does grant on behalf of all authors, a worldwide licence to the Publishers and its licensees in perpetuity, in all forms, formats and media (whether known now or created in the future), to i) publish, reproduce, distribute, display and store the Contribution, ii) translate the Contribution into other languages, create adaptations, reprints, include within collections and create summaries, extracts and/or, abstracts of the Contribution, iii) create any other derivative work(s) based on the Contribution, iv) to exploit all subsidiary rights in the Contribution, v) the inclusion of electronic links from the Contribution to third party material where-ever it may be located; and, vi) licence any third party to do any or all of the above.

