## Supplemental Table 1 for "Genetically predicted serum vitamin D and COVID-19: a Mendelian randomization study"

Supplemental Table 1. Studies contributing to different COVID-19 outcomes in the COVID-19 Host Genetics Initiative

| Study | Percent of COVID-19 outcome sample | | | | |
| --- | --- | --- | --- | --- | --- |
|  | COVID vs. Population | Hosp COVID vs. Population | Sev Resp COVID vs. Population | COVID Pos vs. COVID Neg | Hosp vs. non-hosp COVID |
| Amsterdam UMC COVID study group | 0.11% | 0.16% | 0.24% |  |  |
| Ancestry | 1.25% | 0.23% |  | 13.57% | 20.32% |
| Genetic modifiers for COVID-19 related illness (BelCovid) | 0.11% | 0.16% |  |  |  |
| BoSCO |  | 0.04% | 0.05% |  | 3.68% |
| Biobanque Quebec COVID19 (BQC19) | 0.04% | 0.06% | 0.09% | 0.42% | 1.89% |
| Genetic determinants of COVID-19 complications in the Brazilian population | 0.17% | 0.25% | 0.33% |  |  |
| Genetics of COVID-related Manifestation (Corea) | 0.48% | 0.68% |  |  |  |
| deCODE | 19.82% | 28.30% |  | 24.17% | 17.39% |
| Estonian Biobank | 9.98% |  |  | 9.64% |  |
| FinnGen | 17.19% | 24.62% | 38.00% |  | 2.81% |
| GEN-COVID | 0.23% | 0.31% | 0.47% |  |  |
| genomiCC | 0.72% | 1.04% | 1.60% |  |  |
| Genomics England (genomicsengland100kgp) | 4.50% |  |  | 1.43% |  |
| Genes & Health (GNH) | 1.97% | 2.83% |  | 0.29% | 0.94% |
| Helix Exome+ COVID-19 Phenotypes | 0.40% |  |  |  |  |
| COVID19-Host(a)ge | 0.27% | 0.39% |  |  |  |
| UK Blood Donors Cohort | 3.01% |  |  | 1.00% |  |
| Italy COVID19-Host(a)ge |  |  | 0.31% |  |  |
| Lifelines | 1.84% |  |  | 1.26% |  |
| Michigan Genomics Initiative | 3.71% |  |  | 0.49% |  |
| Million Veterans Program | 1.40% | 0.56% |  | 32.96% | 29.69% |
| Netherlands Twin Register | 0.39% |  |  | 0.20% |  |
| Partners Healthcare Biobank | 2.53% |  |  | 3.16% |  |
| Penn Medicine Biobank | 0.62% | 0.89% |  | 0.86% | 1.52% |
| Qatar Genome Program | 1.01% | 1.38% |  |  | 6.42% |
| Spain COVID19-Host(a)ge |  |  | 0.20% |  |  |
| Determining the Molecular Pathways and Genetic Predisposition of the Acute Inflammatory Process Caused by SARS-CoV-2 | 0.05% | 0.06% | 0.06% |  | 3.32% |
| Genomic epidemiology of SARS-Cov-2 and host genetics in Coronavirus Disease 2019 (COVID-19) | 0.02% |  |  |  |  |
| The genetic predisposition to severe COVID-19 | 0.28% | 0.40% | 0.61% |  |  |
| UK Biobank | 27.87% | 37.65% | 58.05% | 12.28% | 12.01% |
