## Supplemental Table 2 for "Genetically predicted serum vitamin D and COVID-19: a Mendelian randomization study"

Supplemental Table 2. Characteristics of SNPs included in Instrument C

| Gene | SNP | EA | NEA | EAF | BETA | P-VAL | F STAT |
| --- | --- | --- | --- | --- | --- | --- | --- |
| GC | rs11723621 | G | A | 0.29 | -0.19 | 2.9E-1689 | 1467 |
| CYP2R1 | rs10832289 | T | A | 0.41 | -0.07 | 2.03E-266 | 231 |
| NADSYN/DHCR7 | rs12803256 | G | A | 0.78 | 0.10 | 1.3E-378 | 354 |
| CYP24A1 | rs6127099 | T | A | 0.28 | -0.04 | 9.30E-62 | 55 |
| SEC23A | rs8018720 | C | G | 0.82 | -0.03 | 4.04E-36 | 30 |
| AMDHD1/HAL | rs10859995 | C | T | 0.58 | -0.04 | 7.03E-89 | 76 |
| RER1 | rs6698680 | G | A | 0.46 | -0.01 | 8.99E-10 | 7 |
| PADI1 | rs3750296 | C | G | 0.34 | -0.02 | 2.09E-24 | 19 |
| RP4-657M3.2 | rs7519574 | A | G | 0.18 | 0.02 | 2.09E-11 | 9 |
| FOXO6 | rs56044892 | T | C | 0.21 | 0.02 | 2.85E-10 | 8 |
| DOCK7 | rs2934744 | A | C | 0.64 | -0.02 | 3.96E-26 | 23 |
| CELSR2 | rs7528419 | G | A | 0.22 | 0.02 | 2.41E-16 | 13 |
| ARNT | rs3768013 | A | G | 0.37 | -0.01 | 1.37E-13 | 10 |
| FLG | rs61816761 | A | G | 0.02 | 0.13 | 8.57E-74 | 71 |
| FDPS | rs11264360 | A | T | 0.24 | 0.02 | 3.34E-15 | 12 |
| MARC_1 | rs867772 | G | A | 0.68 | -0.01 | 3.64E-11 | 8 |
| -NA- | rs10127775 | T | A | 0.60 | 0.01 | 3.43E-09 | 7 |
| TDRD15 | rs12997242 | A | G | 0.44 | -0.01 | 2.23E-10 | 8 |
| GCKR | rs11127048 | A | G | 0.62 | 0.02 | 6.41E-19 | 16 |
| NPAS2 | rs6724965 | G | A | 0.17 | -0.02 | 1.29E-10 | 8 |
| HTR5BP | rs7569755 | A | G | 0.29 | 0.01 | 8.03E-11 | 8 |
| CPS1 | rs1047891 | A | C | 0.32 | -0.01 | 1.16E-11 | 9 |
| UGT1A4 | rs2011425 | G | T | 0.08 | -0.05 | 9.66E-38 | 31 |
| RHOA | rs7650253 | A | T | 0.69 | 0.01 | 1.76E-10 | 9 |
| CADM2 | rs1972994 | T | A | 0.65 | -0.02 | 7.99E-18 | 14 |
| MRPL3 | rs6438900 | G | C | 0.26 | 0.01 | 9.59E-10 | 7 |
| TFDP2 | rs6773343 | T | C | 0.72 | 0.01 | 5.20E-09 | 6 |
| DOK7 | rs78649910 | A | T | 0.11 | -0.02 | 4.32E-09 | 7 |
| UGT2B7 | rs7699711 | T | G | 0.45 | -0.03 | 6.97E-49 | 41 |
| HSD17B11 | rs58073039 | G | A | 0.30 | -0.01 | 2.16E-11 | 8 |
| ADH1A | rs28364331 | G | A | 0.02 | 0.06 | 1.31E-17 | 14 |
| TNFAIP8 | rs7718395 | G | C | 0.32 | 0.01 | 1.67E-09 | 7 |
| MED23 | rs3822868 | G | A | 0.84 | 0.02 | 1.41E-15 | 13 |
| DNAH11 | rs111529171 | C | G | 0.22 | -0.02 | 6.24E-11 | 8 |
| LINC01004 | rs1011468 | A | G | 0.48 | -0.01 | 1.35E-12 | 9 |
| COG5 | rs1858889 | C | A | 0.50 | 0.01 | 3.85E-11 | 8 |
| GATA4 | rs804280 | A | C | 0.58 | 0.01 | 4.43E-11 | 8 |
| EBF2 | rs34726834 | T | C | 0.25 | 0.01 | 6.65E-10 | 7 |
| LINC00536 | rs7828742 | G | A | 0.60 | -0.02 | 3.06E-28 | 23 |
| DNAH11 | rs10818769 | G | C | 0.86 | -0.02 | 3.35E-09 | 7 |
| ABO | rs532436 | A | G | 0.18 | -0.02 | 2.17E-09 | 7 |
| MAT1A | rs10887718 | T | C | 0.53 | -0.01 | 1.44E-10 | 8 |
| PLEKHA7 | rs567415847 | G | A | 1.00 | 0.28 | 1.03E-14 | 32 |
| TMEM151A | rs523583 | C | A | 0.47 | 0.01 | 5.58E-10 | 7 |
| RP11-21L23.4 | rs1149605 | C | T | 0.17 | 0.02 | 7.34E-14 | 11 |
| ZPR1 | rs964184 | C | G | 0.86 | 0.04 | 5.11E-44 | 37 |
| SLCO1B1 | rs12317268 | G | A | 0.15 | -0.02 | 9.15E-12 | 9 |
| FAM166AP9 | rs9668081 | T | C | 0.47 | 0.01 | 5.38E-09 | 7 |
| LIPC | rs1800588 | T | C | 0.21 | -0.03 | 2.65E-36 | 30 |
| AC007950.2 | rs17765311 | C | A | 0.34 | -0.02 | 1.35E-13 | 10 |
| PEAK1 | rs62007299 | A | G | 0.71 | -0.01 | 1.69E-11 | 9 |
| BCAR4 | rs8063706 | T | A | 0.27 | 0.01 | 3.64E-09 | 7 |
| PDILT | rs77924615 | A | G | 0.20 | -0.02 | 1.46E-10 | 8 |
| FBXL19 | rs71383766 | T | C | 0.42 | 0.01 | 1.15E-09 | 8 |
| CETP | rs1800775 | A | C | 0.49 | -0.02 | 1.56E-17 | 14 |
| RP11-120M18.2 | rs2909218 | T | C | 0.79 | 0.02 | 2.81E-12 | 9 |
| DSG1 | rs8091117 | A | C | 0.07 | -0.02 | 1.03E-09 | 7 |
| SERPINB11 | rs2037511 | A | G | 0.17 | 0.02 | 9.29E-10 | 7 |
| STAP2 | rs57631352 | G | A | 0.30 | -0.01 | 1.48E-09 | 7 |
| LDLR | rs73015021 | G | A | 0.12 | 0.02 | 1.15E-14 | 11 |
| TM6SF2 | rs58542926 | T | C | 0.08 | 0.03 | 8.57E-19 | 15 |
| NPHS1 | rs3814995 | T | C | 0.31 | -0.01 | 2.83E-12 | 9 |
| APOC1 | rs157595 | G | A | 0.61 | -0.02 | 2.95E-14 | 12 |
| SULT2A1 | rs112285002 | T | C | 0.16 | 0.06 | 1.77E-110 | 98 |
| KLK10 | rs10426 | A | G | 0.21 | 0.03 | 3.31E-26 | 21 |
| ZNF808 | rs8103262 | C | T | 0.31 | 0.01 | 3.18E-09 | 7 |
| NRIP1 | rs2229742 | C | G | 0.10 | -0.03 | 7.13E-16 | 12 |
| PLA2G3 | rs2074735 | C | G | 0.06 | 0.03 | 6.55E-12 | 9 |
| SCUBE1 | rs960596 | T | C | 0.34 | 0.01 | 2.23E-09 | 7 |
