## Supplemental Table 3 for "Genetically predicted serum vitamin D and COVID-19: a Mendelian randomization study"

Supplemental Table 3. Minimum detectable odds ratios for COVID-19 outcomes

| COVID-19 outcome | N cases | N controls | Case prevalence | Min detectable  Odds Ratio* |
| --- | --- | --- | --- | --- |
| COVID vs population | 17,965 | 1,370,547 | 0.013 | 0.88 |
| Hosp COVID vs pop | 7,885 | 961,804 | 0.008 | 0.82 |
| Severe resp COVID vs pop | 4,336 | 623,902 | 0.007 | 0.73 |
| COVID pos vs COVID neg | 110,85 | 116,794 | 0.087 | 0.84 |
| Hosp COVID vs non-hosp COVID | 2,430 | 8,478 | 0.223 | 0.66 |

*Minimum detectable odds ratios for 80% power, represents odds ratio per SD increase in serum vitamin D, assumes 3% variance in vitamin D explained by genetic instruments
