## Supplemental Table 4 for "Genetically predicted serum vitamin D and COVID-19: a Mendelian randomization study"

Supplemental Table 4. Replication of MR estimates of effect of vitamin D on COVID-19 outcomes using SNP-vitamin D association data from SUNLIGHT Consortia

| MR method | COVID vs. population | | Hosp COVID vs. population | | Severe Resp COVID vs. population | | COVID pos vs. COVID neg | | Hosp COVID vs. non-hosp COVID | |
| --- | --- | --- | --- | --- | --- | --- | --- | --- | --- | --- |
|  | OR | 95% CI | OR | 95% CI | OR | 95% CI | OR | 95% CI | OR | 95% CI |
| Instrument A | | | | | | | | | | |
| IVW | 1.04 | 0.79-1.35 | 1.04 | 0.67-1.62 | 0.72 | 0.40-1.29 | 1.16 | 0.77-1.75 | 2.39 | 0.93-6.13 |
| MR-Egger intercept test | p-value = 0.90 | | p-value = 0.91 | | p-value = 0.40 | | p-value = 0.26 | | p-value = 0.13 | |

OR, odds ratios per unit increase in serum vitamin D in natural log scale; 95% CI, 95% confidence interval
