## Supplemental Figures 1 - 5 for "Genetically predicted serum vitamin D and COVID-19: a Mendelian randomization study"

**Supplemental Figure 1) Genetic Loci integral to the vitamin D metabolic pathway**

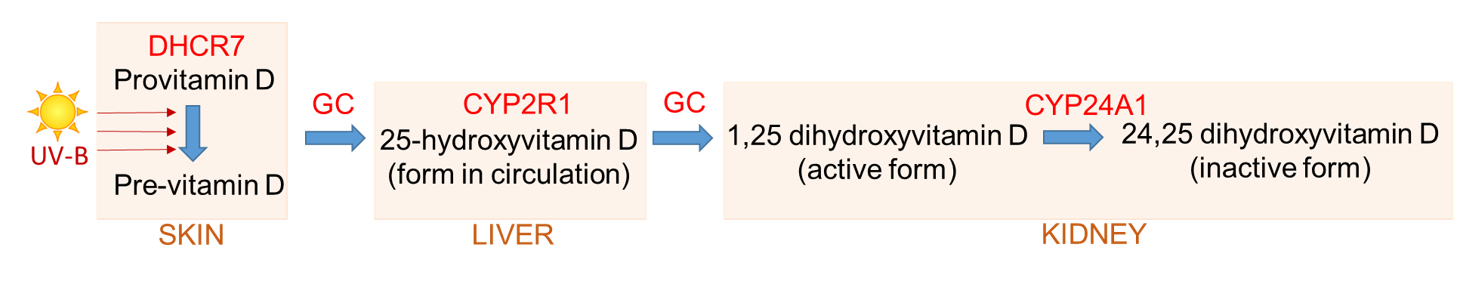

**Supplemental Figure 2) MR scatter and MR forest plots for COVID-19 outcomes**

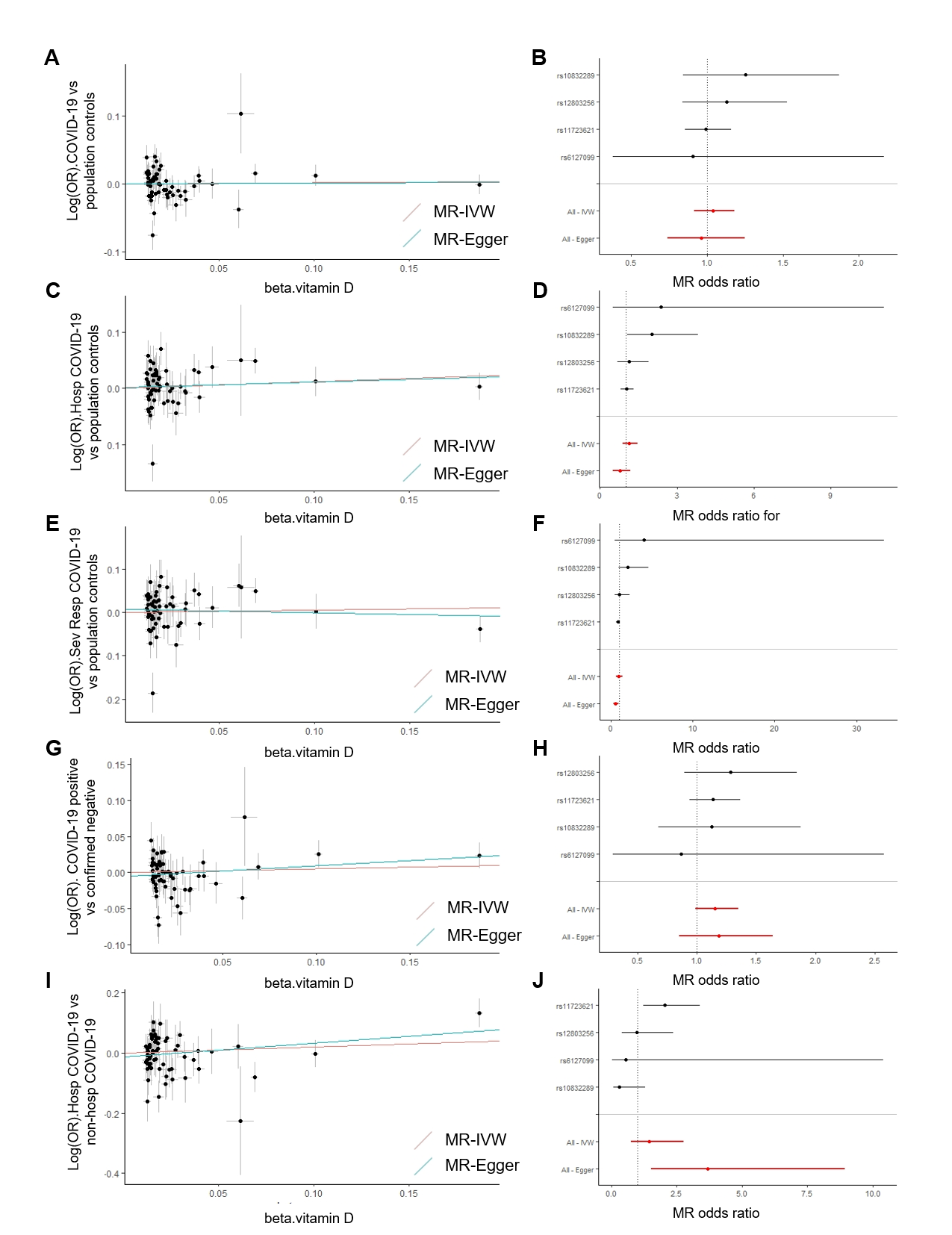

*Supplemental Figure 2 caption: MR scatter and forest plots for (A-B) COVID-19 vs population controls, (C-D) Hospitalized COVID-19 vs population controls, (E-F) Severe respiratory COVID-19 vs population controls, (G-H) COVID-19 vs confirmed negative and (I-J) hospitalized COVID-19 vs non-hospitalized COVID-19. Scatter plots show the SNP—vitamin D associations on the x-axis vs the SNP—log(OR) for COVID-19 case vs comparator on the y axis for all SNPs considered in instruments A,B and C. The MR estimates generated from the IVW (red slope) and MR-Egger (teal slope) MR methods are overlaid. The forest plots show the MR estimates (odds ratios and 95% confidence intervals) generated from instrument A for the effect of 1 SD higher serum vitamin D on risk of COVID-19 case vs comparator. Estimates for each SNP are shown separately (black bars), as well as summarized estimates across all SNPs (red bars).*

**Supplemental Figure 3) Inverse variance weighted MR estimates of effect of vitamin D on risk of COVID-19 infection and hospitalization in European ancestry participants**

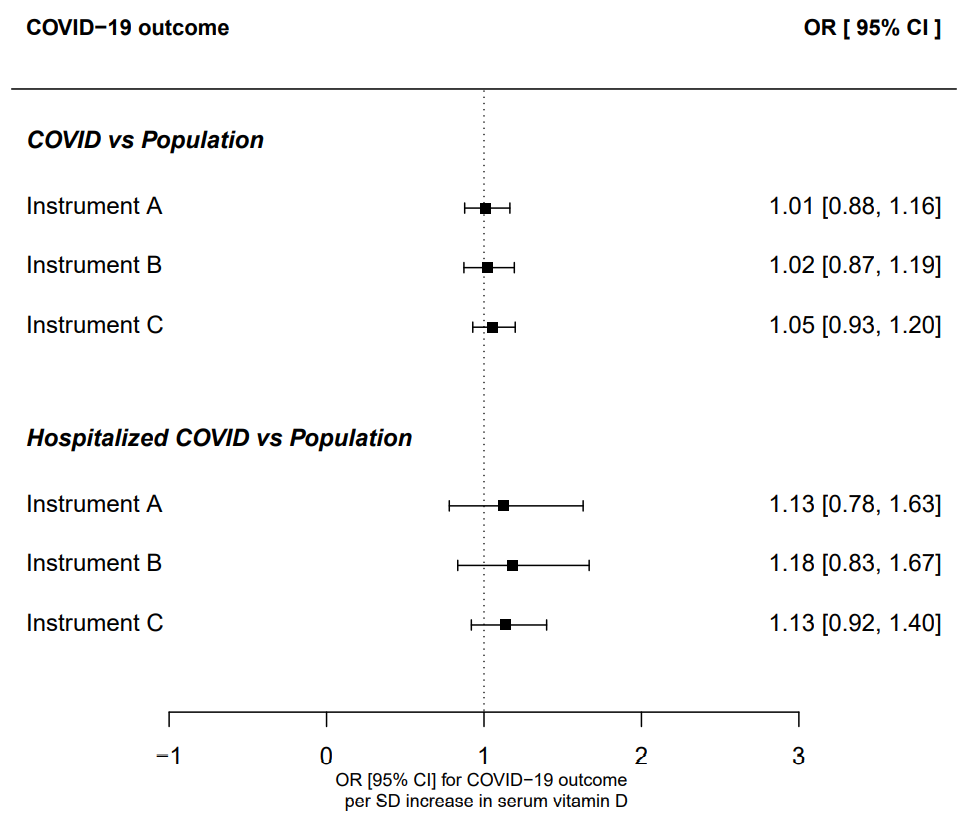

**Supplemental Figure 4) MR estimates of effect of vitamin D on COVID-19 outcomes using different MR Methods for instrument A**

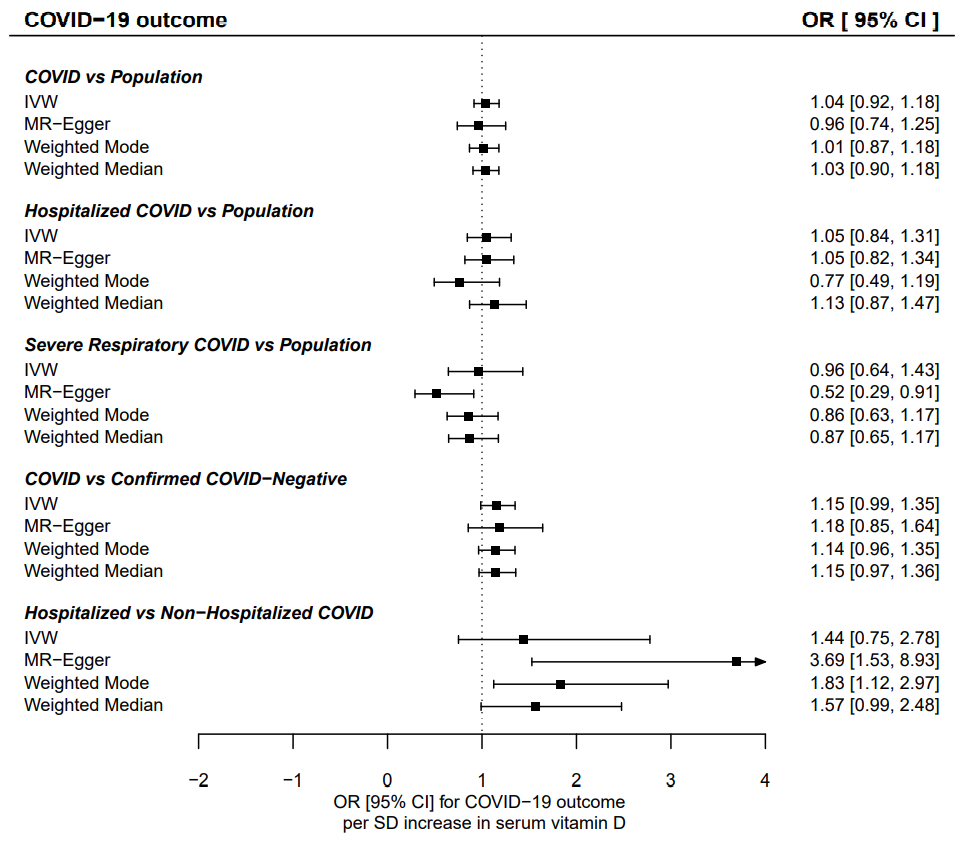

**Supplemental Figure 5) Pairwise correlations of beta-coefficients observed for SNP-vitamin D associations stratified by COVID-19 risk subgroup, across all SNPs in instruments A, B and C**

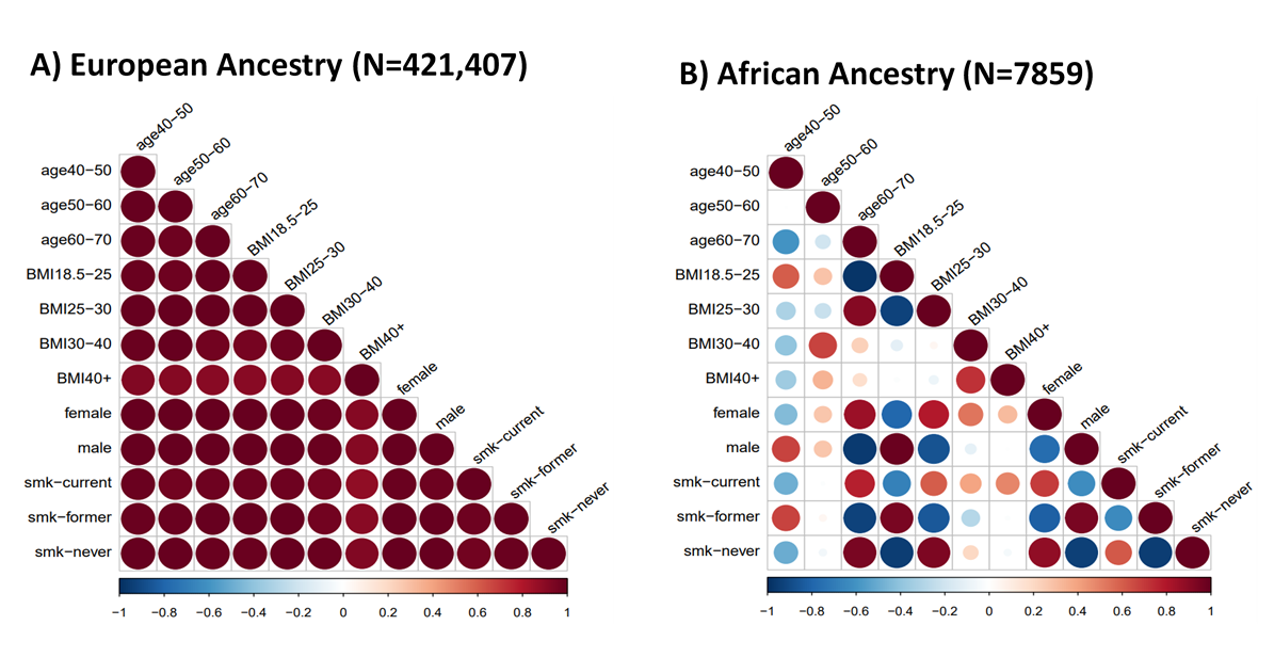
