## Supplementary material for "Genetically predicted serum vitamin D and COVID-19: a Mendelian randomization study": Reporting Checklist

**STROBE-MR checklist**

| Title and abstract  *Indicate Mendelian randomization as the study’s design in the title and/or the abstract.* | Title (line 1); abstract, design and results sections (lines 23 and 35) |
| --- | --- |
| **Introduction** | |
| Background  *Explain the scientific background and rationale for the reported study. Is causality between exposure and outcome plausible? Justify why MR is a helpful method to address the study question.* | Lines 62-99, 95-108 |
| Objectives  *State specific objectives clearly, including pre-specified causal hypotheses (if any).* | Lines 112-114 |
| **Methods** | |
| Study design and data sources  *a) Describe the study design and the underlying population from which it was drawn.*  *Describe also the setting, locations, and relevant dates, including periods of recruitment,*  *exposure, follow-up, and data collection, if available.*  *b) Give the eligibility criteria, and the sources and methods of selection of participants.*  *c) Explain how the analyzed sample size was arrived at.*  *d) Describe measurement, quality and selection of genetic variants.*  *e) For each exposure, outcome and other relevant variables, describe methods of assessment*  *and, in the case of diseases, the diagnostic criteria used.*  *f) Provide details of ethics committee approval and participant informed consent, if relevant.* | a, b and c) Lines 117-129; 154-162; 188-195  d) Lines 131-152  e) Lines 154-174; lines 198-202  f) Lines 226-230 |
| Assumptions  *Explicitly state assumptions for the main analysis (e.g. relevance, exclusion, independence, homogeneity) as well assumptions for any additional or sensitivity analysis.* | Lines 102-105 and 220-224 |
| Statistical methods: main analysis  a) Describe how quantitative variables were handled in the analyses (i.e., scale, units,  model).  b) Describe the process for identifying genetic variants and weights to be included in the  analyses (i.e, independence and model). Consider a flow diagram.  c) Describe the MR estimator, e.g. two-stage least squares, Wald ratio, and related statistics.  Detail the included covariates and, in case of two-sample MR, whether the same  covariate set was used for adjustment in the two samples.  d) Explain how missing data were addressed.  e) If applicable, say how multiple testing was dealt with. | a) Lines 163-174 and 236-238  b) Lines 131-146  c) Lines 240-243  d) NA  e) NA |
| Assessment of assumptions  *Describe any methods used to assess the assumptions or justify their validity.* | Lines 131-152, 176-186, 224-230, 243-245 |
| Sensitivity analyses  *Describe any sensitivity analyses or additional analyses performed.* | Lines 147-152, 212-218, 224-230, 243-245, 259-264, |
| Software and pre-registration  a) Name statistical software and package(s), including version and settings used.  b) State whether the study protocol and details were pre-registered (as well as when and where). | a) Lines 233-236, 257-258, 265-266  b) NA |
| Descriptive data  *a) Report the numbers of individuals at each stage of included studies and reasons for exclusion. Consider use of a flow-diagram.*  *b) Report summary statistics for phenotypic exposure(s), outcome(s) and other relevant variables (e.g. means, standard deviations, proportions).*  *c) If the data sources include meta-analyses of previous studies, provide the number of studies, their reported ancestry, if available, and assessments of heterogeneity across these studies. Consider using a supplementary table for each data source.*  *d) For two-sample Mendelian randomization:*  *i. Provide information on the similarity of the genetic variant-exposure associations between the exposure and outcome samples.*  *ii. Provide information on extent of sample overlap between the exposure and outcome data sources.* | a) Lines 154-162, 289-294, Table 1 b) Supplemental table 3 for outcome prevalence c) Lines 154-162, 188-195, 289-294, Table 1, Supplemental Table 1 d) Lines 328-331, Supplemental Table 1 |
| Main results  *a) Report the associations between genetic variant and exposure, and between genetic variant and outcome, preferably on an interpretable scale (e.g. comparing 25th and 75^th^ percentile of allele count or genetic risk score, if individual-level data available).*  *b) Report causal effect estimate between exposure and outcome, and the measures of uncertainty from the MR analysis. Use an intuitive scale, such as odds ratio, or relative risk, per standard deviation difference.*  *c) If relevant, consider translating estimates of relative risk into absolute risk for ameaningful time-period.*  *d) Consider any plots to visualize results (e.g. forest plot, scatterplot of associations between*  *genetic variants and outcome versus between genetic variants and exposure).* | a) Table 1, Supplemental Table 1, Supplemental Figure 2 b) Lines 308-314, Table 3 c) NA d) Figures 2 and 3, Supplemental Figure 2 |
| Assessment of assumptions  *a) Assess the validity of the assumptions.*  *b) Report any additional statistics (e.g., assessments of heterogeneity, such as I2, Q statistic).* | a) Lines 279-287, 304-306, 314-315, 318-327 |
| Sensitivity and additional analyses  *a) Use sensitivity analyses to assess the robustness of the main results to violations of the assumptions.*  *b) Report results from other sensitivity analyses (e.g., replication study with different dataset, analyses of subgroups, validation of instrument(s), simulations, etc.).*  *c) Report any assessment of direction of causality (e.g., bidirectional MR).*  *d) When relevant, report and compare with estimates from non-MR analyses.*  *e) Consider any additional plots to visualize results (e.g., leave-one-out analyses).* | a) Lines 334-338, Supplemental figure 4  b) lines 293-299, 340-344, Table 3, Figure 3, Supplemental Table 4  c) NA  d) Lines 295-300- expand with data and citation? |
| Key results  *Summarize key results with reference to study objectives.* | Lines 361-372 |
| Limitations  *Discuss limitations of the study, taking into account the validity of the MR assumptions, other sources of potential bias, and imprecision. Discuss both direction and magnitude of any potential*  *bias, and any efforts to address them.* | Lines 409-417, 418-442, 443-453 |
| Interpretation  *a) Give a cautious overall interpretation of results considering objectives and limitations. Compare with results from other relevant studies.*  *b) Discuss underlying biological mechanisms that could be modelled by using the genetic variants to assess the relationship between the exposure and the outcome.*  *c) Discuss whether the results have clinical or policy relevance, and whether interventions could have the same size effect.* | a) Lines 373-385, 386-397  b) Lines 381-383  c) Lines 443-453 |
| *Generalizability*  *Discuss the generalizability of the study results (a) to other populations (i.e. external validity),*  *(b) across other exposure periods/timings, and (c) across other levels of exposure.* | a) Lines 418-442  b and c) Lines 347-359, 443-453, 461-466 |
| Funding | Lines 492-498 |
| Data and data sharing | Lines 509-510 |
| Conflicts of interest | Lines 499-504 |
